# Comparison of Deep Learning Approaches for Conversion of International Classification of Diseases Codes to the Abbreviated Injury Scale

**DOI:** 10.1101/2024.03.06.24303847

**Authors:** Ayush Doshi, Charbel Marche, Pavel Chernyavskiy, George Glass, Thomas Hartka

## Abstract

The injury severity classifications generated from the Abbreviated Injury Scale (AIS) provide information that allows for standardized comparisons in the field of trauma injury research. However, the majority of injuries are coded in International Classification of Diseases (ICD) and lack this severity information. A system to predict injury severity classifications from ICD codes would be beneficial as manually coding in AIS can be time-intensive or even impossible for some retrospective cases. It has been previously shown that the encoder-decoder-based neural machine translation (NMT) model is more accurate than a one-to-one mapping of ICD codes to AIS. The objective of this study is to compare the accuracy of two architectures, feedforward neural networks (FFNN) and NMT, in predicting Injury Severity Score (ISS) and ISS ≥16 classification. Both architectures were tested in direct conversion from ICD codes to ISS score and indirect conversion through AIS for a total of four models. Trauma cases from the U.S. National Trauma Data Bank were used to develop and test the four models as the injuries were coded in both ICD and AIS. 2,031,793 trauma cases from 2017-2018 were used to train and validate the models while 1,091,792 cases from 2019 were used to test and compare them. The results showed that indirect conversion through AIS using an NMT was the most accurate in predicting the exact ISS score, followed by direct conversion with FFNN, direct conversion with NMT, and lastly indirect conversion with FFNN, with statistically significant differences in performance on all pairwise comparisons. The rankings were similar when comparing the accuracy of predicting ISS ≥16 classification, however the differences were smaller. The NMT architecture continues to demonstrate notable accuracy in predicting exact ISS scores, but a simpler FFNN approach may be preferred in specific situations, such as if only ISS ≥16 classification is needed or large-scale computational resources are unavailable.

## 1 INTRODUCTION

Injury severity scores have an important role in trauma injury epidemiology and surveillance research. As part of the Abbreviated Injury Scale (AIS), an anatomy-based coding system developed specifically for trauma injury documentation, the severity designations built into the AIS codes allow for more objective inferences and comparisons to be made for a given set of injuries ^1^. Furthermore, several methods have been proposed to produce a clinically significant, global severity score that encompasses multiple injuries using these severity designations. Two common approaches used to acquire this representative index are the Maximum Abbreviated Injury Scale (MAIS) and the Injury Severity Scale (ISS) methods. The MAIS approach utilizes the highest severity designation for a given set of injuries as the representative index and is frequently used by the National Highway Traffic Safety Administration to evaluate their projects as part of their MAIS-based costs system ^2,3^. The ISS method aggregates the highest severity designations from a maximum of three distinct body regions into a 75-point system and has become the gold-standard for injury severity quantification ^4,5^.

However, the majority of medical data, including visits for trauma injuries, are coded using the International Classification of Diseases (ICD) due to it often being required for billing and reimbursement. Although ICD provides a relatively easy-to-use framework for classifying a wide breadth of diseases and procedures with good inter-operator consistency, a notable downside of this classification system is the lack of associated injury severity designations. This is particularly important to trauma research as it then requires non-standardized inferences to be made about the severity of an injury based on its mechanism and connections with other injury codes ^6^. Given the need for standardized severity metrics for trauma research and that manual AIS coding of injuries for severity designations alone is often prohibitively time-intensive or impossible in some retrospective cases, an automated system to obtain injury severity information using ICD codes is important.

Several methods to facilitate and simplify the conversion of ICD to ISS currently exist. Supported by the organization that maintains AIS, the Association for the Advancement of Automotive Medicine (AAAM), Loftis et al. in 2016 developed the latest official ICD to AIS body region-severity mappings that bridge ICD9 and ICD10 to AIS 2005 with the 2008 update ^7^. In 2010, Clark et al. released a Stata module, ICDPIC, that allowed for the conversion of ICD-9 codes to AIS body region-severity pairs ^8^. The module was then ported into R as ICDPIC-R in 2018 and updated to include ICD-10^9^. More recently in 2023, Hartka et al. developed a neural machine translation-based tool that allowed for the conversion of ICD10 to full AIS 2005 with 2008 update codes using the National Trauma Data Bank ^10^. This NMT model showed improved accuracy compared to the official AAAM map and ICDPIC(-R) methods, especially for major trauma defined by an ISS ≥16. However, these methods generally fall short in several metrics. Studies analyzing the 1:1 translation accuracy of the official AAAM mappings compared to manual coding by certified AIS coders demonstrated moderate accuracy ranging from 70-82% and poor inter-operator agreement as low as 48% ^11–13^. Studies evaluating the accuracy of ISS scores calculated from ICD codes converted using ICDPIC-R demonstrated an overall accuracy of 17.7% ^10,14^. Finally, all three methods of ICD to ISS score conversion globally underestimate the ISS score compared to those calculated from manually-coded AIS codes ^10,13,14^.

The results from the NMT approach indicate that deep learning is a promising technology for ICD to ISS conversion. However, the use of an NMT to first convert ICD codes to AIS codes and then calculate the ISS score from the converted AIS codes may be more complex than necessary. The goal of this study was to compare the performance of the simpler feed-forward neural network (FFNN) architecture to the performance of the more complex NMT architecture in predicting severity information, as well as determine if there is an advantage in direct prediction from ICD codes compared to indirect prediction through AIS.

## 2 METHODS

This study compared two different machine learning architectures, FFNN and NMT, using two different model structures for each, direct conversion from ICD to ISS and indirect conversion through AIS. These models were trained and tested using data from the U.S. National Trauma Data Bank (NTDB).

### 2.1 Datasets

The NTDB is a dataset managed by the American College of Surgeons that contains trauma injury cases reported from every Level-I, Level-II, and Level-III trauma center in the United States as part of their credentialing requirement. Specifically, the dataset covers trauma injury cases where the patient either was admitted to the hospital or died in the emergency department. Although Level-IV, Level-V, and community hospitals are not required to report injury cases, they are able to do so voluntarily.

Every case in the NTDB contains demographic information about the patient, the mechanism of initial injury, procedures, diagnoses, and outcomes. The patient’s age, sex, and co-morbidities are included in the demographics. The initial mechanism of injury is reported using ICD external-cause codes (E-codes) while any procedures that were performed are reported using ICD procedures codes (P-codes). The diagnoses were doubled coded in both ICD and AIS manually by trained registrars certified in both systems. Double manual coding provides the current gold-standard data for ICD to AIS and thereby ICD to ISS conversions. It has been used to test other conversion models, including AAAM’s official ICD to AIS mappings ^10,13^.

NTDB data from 2017-2018 was used to train and validate the models while data from 2019 was used to test them. During these selected years, the diagnoses for a given case were reported using both ICD-10 and AIS 2005 with 2008 update. The 2,031,793 trauma cases from 2017-2018 were pooled together and randomly assigned to training and validation datasets at a 90%-10% ratio, resulting in 1,828,613 being used for training and 203,180 cases being used for validation. The testing dataset comprised all 1,091,792 trauma cases from 2019. Data from a separate year was chosen for testing to account for any minor inter-year changes in coding practices.

### 2.2 Outcomes of interest

The two primary outcomes of interest for this study were exact ISS score prediction and ISS ≥16 classification accuracy. ISS ≥16 is a commonly used cutoff for classifying a patient as severely injured based on mortality rates identified in the Major Trauma Outcome Study ^15,16^. The gold standards used to assess model performance were the ISS score and ISS ≥16 classification calculated from the corresponding manually coded AIS codes. Sensitivity and specificity were reported alongside ISS ≥16 classification prediction accuracy due to it being an imbalanced metric within the dataset with most cases failing to meet criteria. Further subpopulation sensitivity and specificity analysis were performed on ISS ≥16 classification performance stratified by sex and age.

The secondary outcomes of interest were MAIS ≥3 classification prediction accuracy and the percent of all predicted AIS codes that were correct. A predicted AIS code was considered correct if it either exactly matched a manually coded AIS code or shared both body region and severity with one. These secondary outcomes were only performed on indirect models given their intermediate AIS prediction step, which is skipped in direct models.

### 2.3 FFNN models

The two FFNNs developed for this study were built using the PyTorch framework, which is an open-source, machine-learning framework based on the tensor library Torch. Both the direct and indirect FFNN models contained a parametric rectified linear unit (PReLU) layer between two linear transformation layers that were initialized using the Kaiming uniform method ^17^. The initial value for the PreLU layer was 0.25. The output function for the multiclass classifier, the direct FFNN, was a LogSoftmax layer while the output function for the multilabel classifier, the indirect FFNN, was a Sigmoid layer. Weights were adjusted using an Adagrad optimizer with an initial *β*_1_ and *β*_2_ of 0.9 and 0.98, respectively. The initial learning rate was 0.01 with a decay factor of 5 for every two consecutive un-improving epochs and an early stop condition of 10 decays. A negative log likelihood loss function was used during training of the direct FFNN while a binary cross entropy loss function was used for the indirect FFNN.

As input for the FFNN models, every age and sex demographic, E-code, P-codes, and ICD-10 diagnosis code present in the dataset were combined and transformed into a binary dummy variable system. Each trauma case was then converted into a sparse binary tensor of these dummy variables, which were used as input for both the direct and indirect models. A similar system was used for the outputs, with every possible ISS score being converted to a dummy variable system for the direct FFNN and every AIS code being used for the indirect FFNN. However, the method for selecting the predictions from the output tensor differed between the direct and indirect structures. The ISS dummy variable with the largest predicted score from the LogSoftmax layer was chosen for the direct structured model while any AIS dummy variable with a predicted score greater than 0.3 from the sigmoid layer was selected for the indirect structured model.

### 2.4 NMT models

The two NMTs developed for this study were built using the PyTorch implementation of OpenNMT, an open-source toolkit developed to research NMTs and perform competitively. The NMT models were based on the Transformer architecture published by the Google Brain team ^18^. Eight attention heads with a dropout of 0.1 were used and the encoder-decoder stacks contained six identical, 512-unit layers. Each encoder layer contained a multi-head self-attention mechanism followed by a position-wise FFNN and layer normalization. Each decoder layer contained a masked multi-head attention mechanism followed by a similar multi-head self-attention and FFNN mechanisms as the encoder layers. Weights were adjusted using an Adam optimizer with an initial *β*_1_ and _2_ of 0.9 and 0.998, respectively, and an initial learning rate of 2. The learning rate decay was proportional to the inverse square root of the step number and a categorical cross-entropy loss function was used for training.

The input for the NMT models were sentences generated by concatenating the age and sex of the patient, the E-code, any P-codes, and the ICD-10 diagnosis codes for each trauma case into a space-separated string without periods. These sentences were used as input for both the direct and indirect NMT models. The age, P-codes, and ICD diagnosis codes were prefixed with an A, P, and D, respectively, to separate them in the vocabulary structure generated by the model as well as increase readability when examining attention results. The output sentences differed between the direct and indirect structures. For the direct structure, the output sentence was only the ISS score. For the indirect structure, the output sentence was a space-separated string of AIS codes without the severity designation arranged in ascending numerical order.

### 2.5 Testing and comparing the models

Predicted ISS scores and ISS ≥16 classifications were generated for the testing dataset using the four models and were compared to expected scores and classifications from the databank. Accuracies for correctly predicting the exact ISS scores and ISS ≥16 classifications were calculated for the four models and compared to one another. The statistical significance of the differences in performance were tested using Cochran’s Q test followed by post-hoc pairwise McNemar tests. Root mean squared error (RMSE) was used to further compare the four models in predicting exact ISS scores while sensitivity and specificity analysis was performed on the ISS ≥16 classification results. Additional subpopulation sensitivity and specificity analysis were performed after stratifying for both sex and age, with age being binned into 0-17, 18-64, and 65+ year groups.

For the secondary outcomes, the predicted MAIS ≥3 classifications for the two indirect models were compared against the expected MAIS ≥3 classifications. The statistical significance of the difference in performance was tested through a McNemar test. The quality of the AIS code predictions were analyzed by calculating the percentage of predicted codes that either exactly matched or shared the same body region and severity with an expected code. This percentage was calculated from the union set of both predicted and expected codes for each case. Statistical differences between the two sets of percentages were then compared using the Wilcoxon signed-rank test and effect size was reported as the pseudo-median difference.

## 3 RESULTS

### 3.1 Model training and testing

The demographic and injury statistics for the training, validation, and testing datasets are shown in Table 1. The number of injuries per patient, the distribution of ISS scores, and the percentage of MAIS ≥3 classifications were similar across the three datasets. However, the testing dataset had a slightly older population with more incidence of falls, less male predominance, and a lower prevalence of ISS ≥16 classifications compared to the training and validation datasets. Yet, given that the testing dataset was obtained from a different year to allow for robustness testing against inter-year differences, some variation in the distributions was expected.

**Table 1:** Demographic and injury statistics of the patients in the training, validation, and testing datasets.

|  | Training Dataset | Validation Dataset | Testing Dataset |
| --- | --- | --- | --- |
| Years | 2017 - 2018 |  | 2019 |
| Total number of patients | 1,828,613 | 203,180 | 1,091,792 |
| Number of injuries per patient (Median [IQR]) | 2 [1-4] | 2 [1-4] | 2 [1-4] |
| Age in years (Median [IQR]) | 47 [23-68] | 47 [23-68] | 49 [24-70] |
| Males (Percentage) | 59.71% | 59.61% | 58.90% |
| Mechanism of Injury (Percentage) |  |  |  |
| Falls | 46.6% | 46.5% | 49.2% |
| Automotive-related | 32.2% | 32.2% | 30.3% |
| Assault | 9.2% | 9.2% | 8.5% |
| Self-injury | 1.3% | 1.2% | 1.4% |
| Other | 10.7% | 10.7% | 10.6% |
| ISS (Median [IQR]) | 8 [4-10] | 8 [4-10] | 8 [4-10] |
| ISS $\geq 16$ (Percentage) | 15.8% | 15.7% | 15.2% |
| MAIS $\geq 3$ (Percentage) | 31.0% | 31.1% | 31.3% |

Training and testing durations for the four models are shown in Table 2. Testing was performed twice, once with a CUDA-enabled GPU and once without it. Both training and GPU-inclusive testing were performed on a computing cluster with an allocation of four standard processing nodes, 60 GB of RAM, and an NVIDIA A100 80GB VRAM Tensor Core GPU. CPU-only testing was performed on a computer cluster with similar four processing nodes but without a GPU allocation. Furthermore, the RAM size requirements differed between the FFNN and NMT models for the CPU-only testing. The FFNN models were able to convert ICD codes with a smaller 8 GB RAM allocation while the NMT models required a larger 32 GB of RAM. Conversion outputs for both GPU-inclusive and CPU-only testing were identical.

**Table 2:** Training and testing durations of the four models.

|  | Computation Times |  |  |  |
| --- | --- | --- | --- | --- |
|  | Direct FFNN | Indirect FFNN | Direct NMT | Indirect NMT |
| Training | 11 hrs., 10 mins | 11 hrs., 22 mins | 12 hrs., 27 mins | 14 hrs., 34 mins |
| Testing (GPU-inclusive) | 1 min, 6 s | 1 min, 24 s | 69 min, 10 s | 82 mins, 53 s |
| Testing (CPU-only) | 62 min, 7 s | 97 min, 2 s | 251 min, 32 s | 307 min, 19 s |

### 3.2 Accuracy in exact ISS score prediction

The first primary outcome of interest was the accuracy of the four models in predicting the exact ISS score. As shown in Table 3, the indirect NMT model continued to perform the best with an accuracy of 79.8%, followed by the direct FFNN (76.5%), direct NMT (75.9%), and indirect FFNN (74.3%) models. Cochran’s Q testing followed by post-hoc pairwise McNemar testing without continuity correction demonstrated that the differences in performance between each pairwise combination were statistically significant (Table 4). When comparing RMSE, the two NMT models demonstrated smaller overall errors than the two FFNN models (Table 3). Furthermore, the relative rankings of the direct FFNN and direct NMT models’ performance using RMSE was discordant with their respective rankings when using accuracy, with the overall less accurate direct NMT model demonstrating smaller average errors than the overall more accurate direct FFNN.

**Table 3:** Performance in exact ISS score prediction for the four tested models. Accuracy was measured by comparing the predicted testing dataset ISS scores for each model to the expected ISS scores in the NTDB. RMSE was calculated for each model using the differences between the predicted and expected scores.

|  | Exact ISS Score |  |
| --- | --- | --- |
|  | Accuracy | Root Mean Squared Error |
| Direct (to ISS) FFNN | 76.5% | 4.06 |
| Indirect (to AIS) FFNN | 74.3% | 4.51 |
| Direct (to ISS) NMT | 75.9% | 3.83 |
| Indirect (to AIS) NMT | 79.8% | 3.77 |

**Table 4:**
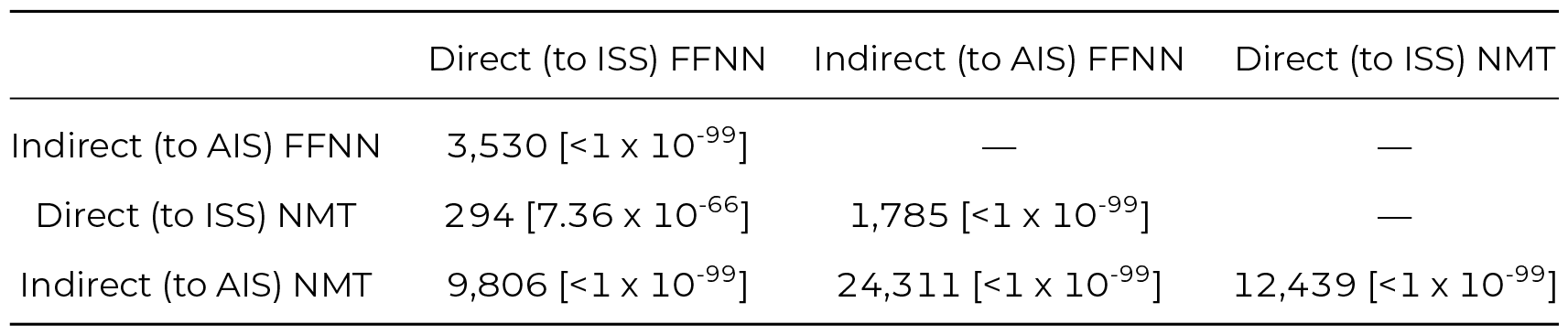
McNemar statistics and associated adjusted *p-values* from each post-hoc pairwise McNemar test on exact ISS score prediction performance. Differences between all pairwise combinations were found to be statistically significant.

### 3.3 Accuracy in ISS ≥16 classification prediction

The secondary primary outcome of interest was the accuracy of the four models in predicting the ISS ≥16 classification. Similar rankings were seen in ISS ≥16 classification performance compared to exact ISS score performance, with the indirect NMT model continuing to have the best accuracy for ISS ≥16 classification (94.0%), followed by the direct FFNN (93.4%), direct NMT (93.1%), and indirect FFNN (93.1%) models (Table 5). Similar Cochran’s Q and post-hoc pairwise McNemar testing demonstrated that the differences in performance between each pairwise combination was statistically significant except for the direct NMT and indirect FFNN comparison (Table 6). On global sensitivity and specificity analysis, all four models had similarly high specificity; however, the NMT models demonstrated superior sensitivity against the FFNN models overall.

**Table 5:** Performance in ISS ≥16 classification for the four tested models. Accuracy, sensitivity, and specificity were measured by comparing the predicted testing dataset ISS ≥16 classifications for each model to the expected ISS ≥16 classifications from the reported ISS scores in the NTDB.

| | ISS $\geq 16$ Classification | | |
| --- | --- | --- | --- |
|  | Accuracy | Sensitivity | Specificity |
| Direct (to ISS) FFNN | 93.4% | 65.1% | 97.6% |
| Indirect (to AIS) FFNN | 93.1% | 66.3% | 97.8% |
| Direct (to ISS) NMT | 93.1% | 72.2% | 96.8% |
| Indirect (to AIS) NMT | 94.0% | 74.9% | 97.4% |

**Table 6:** McNemar statistics and associated adjusted *p-values* from each post-hoc pairwise McNemar test on ISS ≥16 classification performance. Differences between all pairwise combinations were found to be statistically significant except for the direct NMT model and indirect FFNN model comparison.

|  | Direct (to ISS) FFNN | Indirect (to AIS) FFNN | Direct (to ISS) NMT |
| --- | --- | --- | --- |
| Indirect (to AIS) FFNN | 220 [$2.56 \times 10^{-49}$] | — | — |
| Direct (to ISS) NMT | 193 [$1.72 \times 10^{-43}$] | 0.708 [0.4] | — |
| Indirect (to AIS) NMT | 867 [$<1 \times 10^{-99}$] | 1,846 [$<1 \times 10^{-99}$] | 1,608 [$<1 \times 10^{-99}$] |

Table 7 and Table 8 show the results from the subpopulation-stratified sensitivity and specificity analyses on the performance of the four models based on sex and age groupings, respectively. Each of the four models demonstrated consistently similar specificities across all sex and age subpopulations compared to its global specificity. However, large variations were seen across the subpopulations on sensitivity analysis. When stratified by sex, all models had a higher sensitivity for the male subgroup than their global sensitivity along with the respective inverse for the female subgroup (Table 7). The performance rankings of the models on sensitivity mirrored that of the global sensitivity rankings, with the NMT models outperforming the FFNN models. Furthermore, the NMT models demonstrated smaller differences between the male and female subgroups than the FFNN models while the indirect models demonstrated higher sensitivities than their respective direct models. When stratified by age grouping, analogous patterns in the sensitivity variations to that of the sex-stratified analysis was observed. For all models, the 18-64 year group demonstrated the highest sensitivity and was greater than each model’s respective global sensitivity (Table 8). Inversely, both the 0-17 and 65+ year groups underperformed in sensitivity compared to their respective global sensitivity, with the 65+ year group demonstrating the lowest sensitivity for all age-group-stratified subgroups across all models. The NMT models’ sensitivities were overall larger than that of the FFNN models’, which is consistent with patterns seen on the global analysis. Additionally, the indirect models continued to demonstrate higher sensitivities than their respective direct models.

**Table 7:**
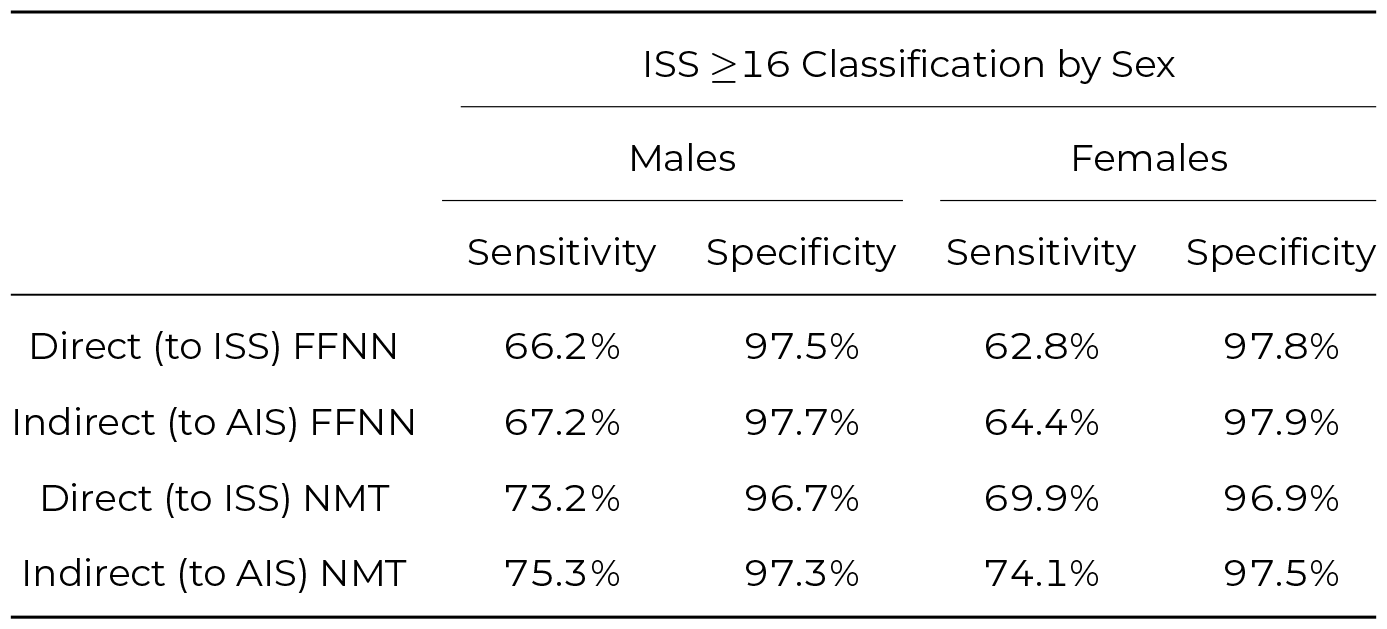
Sex-stratified subpopulation sensitivity and specificity analysis of the ISS ≥16 classification performance for the four models.

**Table 8:** Age-group-stratified subpopulation sensitivity and specificity analysis of the ISS ≥16 classification performance for the four models.

| | ISS $\geq 16$ Classification by Age Group | | | | | |
| --- | --- | --- | --- | --- | --- | --- |
|  | 0 - 17 yrs. |  | 18 - 64 yrs. |  | 65+ yrs. |  |
|  | Sensitivity | Specificity | Sensitivity | Specificity | Sensitivity | Specificity |
| Direct (to ISS) FFNN | 62.2% | 97.9% | 69.2% | 97.4% | 58.1% | 97.7% |
| Indirect (to AIS) FFNN | 62.1% | 98.2% | 69.8% | 97.6% | 60.9% | 97.7% |
| Direct (to ISS) NMT | 68.7% | 97.1% | 76.6% | 96.6% | 64.8% | 96.9% |
| Indirect (to AIS) NMT | 73.3% | 97.6% | 77.1% | 97.5% | 71.1% | 97.1% |

### 3.4 Accuracy in MAIS ≥3 classification and AIS code prediction

The two secondary outcomes of interest were the MAIS ≥3 classification and AIS code prediction accuracy for the two indirect models. The two direct models were not included in these comparisons as they both directly predicted ISS scores without predicting AIS codes as an intermediary step. The indirect NMT model was more accurate than the indirect FFNN model in predicting MAIS ≥3 classifications at 94.0% and 91.8%, respectively (Table 9). McNemar testing demonstrated a statistically significant difference in the accuracy of the two models with a *p-value* of <1 × 10^−99^. Furthermore, the indirect NMT model predicted correct AIS codes at a higher percentage than the indirect FFNN model at 88.3% and 79.6% respectively. Wilcoxon signed rank testing of the two paired sets of percentages demonstrated a statistically significant pseudo-median difference of 21% for the two non-parametric distributions with a *p-value* of <1 × 10^−99^.

**Table 9:**
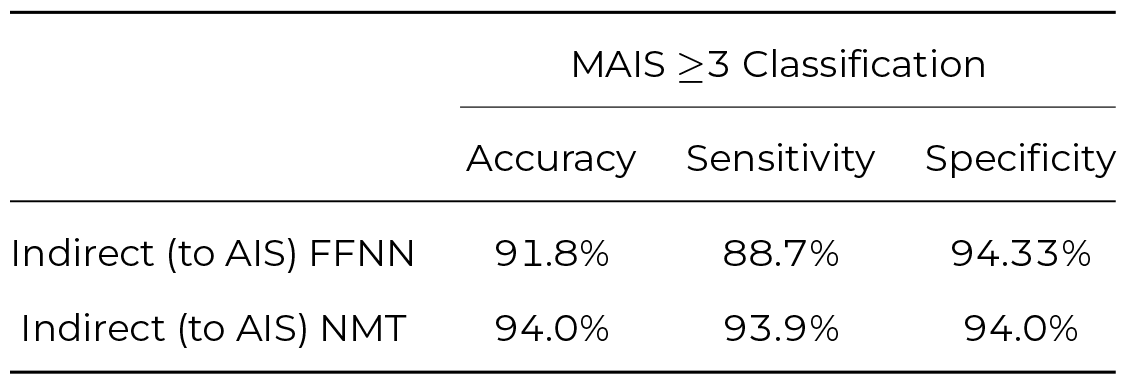
Performance in MAIS ≥3 classification for the two indirect models. Accuracy, sensitivity, and specificity were measured by comparing the predicted testing dataset MAIS ≥3 classifications for each model to the expected MAIS ≥3 classifications from the reported AIS codes in the NTDB.

## 4 DISCUSSION

In this study, three newly proposed machine learning models, direct FFNN, indirect FFNN, and direct NMT, were compared against the previously proposed indirect NMT model in predicting injury severity scores and classifications from ICD-10 codes. The indirect NMT model was found to notably outperform the other models in predicting the exact ISS score, but demonstrated only marginal improvement over them in predicting ISS ≥16 and MAIS ≥3 classifications based on accuracy. Furthermore, the NMT and FFNN architectures demonstrated similarly high specificities in binary classification tests, but the NMT models were more sensitive across the board in those same metrics.

### 4.1 FFNN models vs NMT models

The results of this study demonstrate that there exist different, viable applications of deep learning in acquiring standardized injury severity data from cases only coded using the ICD-10 system. While manual coding by certified experts will continue to remain the gold standard for acquiring injury severity information, the option to generate it in situations where manual AIS coding is either impractical or impossible would be a powerful tool in the field of injury research. Although the previously proposed indirect NMT model provided the most accurate injury information overall, the simpler FFNN models could be used instead in specific situations.

In predicting the exact ISS score and severity classification, both the FFNN and NMT models performed similarly well in terms of accuracy, especially with ISS ≥16 classification. Furthermore, both approaches were equally specific in accurately predicting binary classification results. However, the NMT models generated smaller errors in aggregate compared to the FFNN models based on the RMSE. Similar distinctions are seen with both ISS ≥16 and MAIS ≥3 classification, as the NMT models demonstrated higher sensitivities than the FFNN models, including on subpopulation analysis. Though the exact cause of this stratification is unclear, a potential explanation is that the FFNN approach is not as strongly generating associations during the training process due to data spareness, even when using an appropriate Adagrad optimizer. This results in muddying the decisiveness of the prediction, both in not meeting the 0.3 prediction score cutoff criteria for the multilabel indirect model as well as not generating distinct predictions for the multiclass direct model. On the other hand, the multiple multi-head attention and feedforward layers comprising the encoder-decoder structure of the NMT lends itself to faster and more decisiveness learning from sparse data given its development initially for language translation ^18^. This shortcoming of the FFNN results in an overall underscoring of ISS in the direct FFNN case or predicting fewer AIS codes and thereby underestimating the ISS score in the indirect FFNN case. This distinction is further supported through the comparison of the indirect FFNN and indirect NMT models in their AIS code prediction accuracy, with the indirect NMT significantly outperforming the indirect FFNN.

The other important practical difference between the FFNN and NMT architectures is the significantly shorter conversion time of the FFNN models compared to the NMT models. This difference is understandable given that the FFNN architecture is significantly smaller with only 4 layers compared to the much larger NMT architecture using multiple encoder-decoder units. In situations with limited computational resources with respect to both power and space, the smaller FFNN model may be preferable or even the only viable option. This is demonstrated with the NMT models requiring at least 32 GB of RAM compared to the 8 GB of RAM required by the FFNN models during CPU-only testing. Additionally, although the FFNN models are outperformed by the NMT models on key metrics, both FFNN models outperformed other known options for calculating ISS, namely the official AAAM mapping and ICDPIC-R ^10^.

### 4.2 Direct (ICD to ISS) vs indirect (ICD to AIS to ISS) approaches

Comparison of models’ performance stratified by direct or indirect approach demonstrated a statistically significant, but clinically insignificant, difference. For both the FFNN models and the NMT models, the use of either a direct or indirect approach resulted in similar accuracies, sensitivities, and specificities. Furthermore, no consistency was found in the relative performance of the direct versus the indirect models. The direct approach outperformed the indirect approach when using a FFNN architecture while the inverse was found when using a NMT architecture. The most notable difference between the two approaches arises from computation times, as the direct models were slightly faster in conversion compared to their respective indirect counterparts. However, the reduction in computation time is significantly smaller than the computation time differences between the FFNN and NMT models.

### 4.3 Study strengths

The main strengths of this study were the two-by-two comparisons of architectures and approaches, the large sample size of the datasets, and the robustness of the testing data. By performing a two-by-two comparison using the four different approach-architecture pairs, distinctions and inferences could be made about the effects the model architecture or approach independently had on the accuracy of its estimations. Additionally, the large sample size from the NTDB provided enough cases to both adequately train the models and help counteract the effects of each individual code’s relative sparseness in the database. Furthermore, the large sample size of testing dataset helped increased the power of the study. Finally, by using a different year for the testing dataset compared to the training dataset, the models’ robustness against expected, small inter-year variability in coding practice would also be assessed.

### 4.4 Study limitations

There are several important limitations to consider for this study. First, it is unknown whether the layers and structures used for the models are the most optimal. Given both the stochastic nature of training a deep learning model as well as the variety of layers, activation functions, and sizes that can be combinatorially used, it is impossible to know where the true global maximum in performance lies. Furthermore, the cutoff used for the indirect FFNN of 0.3 was chosen through analysis of the model’s performance on the testing dataset with the goal of maximizing accuracy. However, this cutoff may vary for different patient population as the *a priori* probability of a given AIS code will be different. Second, the limitations of the NTDB are propagated into this study from its use. Namely, given the higher acuity population that constitutes the NTDB due to how cases are submitted, it is unknown how the differences in model performance will change in the setting of a lower acuity population. Further validation of these models will be needed to better understand the generalizability of these approaches.

## 5 CONCLUSIONS

A variety of deep learning model architectures and approaches can be used in the estimation of injury severity with varying levels of accuracy when the resources or data for manual coding with AIS is unavailable. The indirect NMT model demonstrated the best performance compared to the other three models overall; however, the other three models demonstrated similar efficacy in specific situations, namely for binary severity classifications with limited computational resources. In these situations, a less computationally intense and faster model may be preferable, especially for the conversions of larger datasets.

## Data Availability

All data produced in the present study are available upon reasonable request to the authors.

## 6 ACKNOWLEDGEMENTS

The authors would like to thank the National Trauma Data Bank (NTDB) for providing the data to perform this study. The content reproduced from the NTDB remains the full and exclusive copyrighted property of the American College of Surgeons. The American College of Surgeons is not responsible for any claims arising from works based on the original data, text, tables, or figures.

